# An atlas of exposome-phenome associations in health and disease risk

**DOI:** 10.1101/2025.06.05.25329055

**Authors:** Chirag J Patel, John PA Ioannidis, Arjun K Manrai

## Abstract

Non-genetic exposures comprising the “exposome,” including diet, lifestyle, infections, and pollutants, shape many clinical phenotypes, yet the evidence remains fragmented. We conducted an Exposome-Wide Association study (ExWAS) incorporating 619 exposure indicators and 305 quantitative phenotypes across 10 independent waves of the US Centers for Disease Control and Prevention National Health and Nutrition Examination Survey (CDC NHANES). Replicable and stable signals were most concentrated in cardiometabolic and anthropometric phenotypes, linking objective nutrient biomarkers and lipophilic pollutants with body mass index, glycated hemoglobin, and lipid profiles. Triglycerides, an important marker for cardiovascular risk, emerged as the phenotype most strongly associated with multi-domain exposures, notably trans fatty acids, persistent pollutants, and vitamin E isoforms. In pulmonary traits, tobacco-specific and carcinogen biomarkers were more prominently associated with reduced lung function than short-lived nicotine metabolites, refining exposomic links to Forced Expiratory Volume in 1 second (FEV1). While individual exposures showed modest effects, aggregate “poly-exposomic” models explained phenotypic variation comparable to genome-wide polygenic scores. Exposome globes further reveal an interconnected architecture where exposures rarely act in isolation, complicating causal attribution while providing a more holistic view of environmental risk. Our findings highlight which exposures are most likely to add value to disease risk assessment, population surveillance, as well as further exposure prioritization and next-generation longitudinal exposomics.

## Introduction

Clinically-relevant phenotypes are influenced by both genetics and environmental exposures^1–3^. Despite this, the structural relationship between the exposome– defined as the totality of environmental exposures in broad physical, chemical, and psychosocial domains^4,5^—and human health remains obscure, characterized by a lack of systematic mapping across its broad domains. To date, interrogating exposome-phenome relationships has been limited to studies that target a few candidate exposures and phenotypes. These candidate studies are presented selectively in millions of papers on claimed associations yielding fragmented and often biased snapshots of the exposome-phenotype maze ^6^. While candidate approaches have been successful in identifying factors with large effects, such as smoking^7^, these millions of studies to-date have not yielded robust associations ^8^ moreover, many reported results might be false positives ^9^. For example, disciplines such as nutritional epidemiology have yielded numerous associations regarding single dietary factors and patterns in disease outcomes have been non-robust^10,11^. Analogous debates have been made in fields studying other domains of the exposome, such as environmental epidemiology ^12,13^. Previous criteria to gauge causality, such as those developed by Bradford and Hill ^7^ may not be readily applicable in new exposome epidemiology scenarios, e.g. if most of the true associations to be discovered have small effect sizes and not readily discernible biological plausibility^14^, analogy, coherence and specificity, and there is no possibility to validate in experimental studies. Consequently, the opportunity to integrate environmental data into precision medicine remains underrealized.

Precision medicine approaches, however, are dominated by genetic factors. Which exposures, if measured, would meaningfully improve risk stratification or refine prognosis, and how large are those effects relative to demographics and genetics? Many phenotypes routinely used for care, diagnosis, staging, and risk prediction, such as lipids (e.g., triglycerides), HbA1c and fasting glucose, eGFR/creatinine, inflammatory markers (e.g., C-Reactive Protein [CRP]), and spirometry (Forced Respiratory Volume in 1 Second [FEVL]), may be partially driven by modifiable exposures. Prioritizing clinical phenotypes by the magnitude and replicability of associations, contextualizing connections between smoking and nutrient biomarkers, and quantifying variance explained to gauge utility for risk equations are needed for evaluating the role of the exposome in precision medicine.

Here, we hypothesize that the exposome exhibits a replicable associational architecture where aggregate factors explain clinically relevant phenotypic variance and disease risk. To evaluate this, we systematically quantify these relationships, executing an “exposome-wide association study”, establishing the data-driven foundation required to integrate the exposome into precision medicine ^15^.

## Results

In brief, we developed an analytic pipeline (Figure 1, Extended Data Figure 1) to analyze data from participants of the National Health and Nutrition Examination Survey (NHANES)^16^ in 10 serial cross-sectional surveys that were sampled in years 1999-2000, 2001-2002, 2003-2004, 2005-2006, 2007-2008, 2009-2010, 2011-2012, 2013-2014, 2015-2016, and 2017-2018. We cataloged a total of 374 real-valued continuous phenotypes and 810 biomarkers or self-reported questionnaires responses that measure pollutant, dietary, infectious, or smoking-related exposures across all 10 surveys. See Supplementary Results for sample size and power. Figure S1 describes the distribution of demographic characteristics (sex, age, ethnicity, education, income) for each association. The median age was 40 (interquartile range [IQR]: 34, 42) and the median income-to-poverty ratio was 2.9 (IQR: 2.8, 2.9). All associations are found in Table S1. The exposure and phenotype catalogs are available in Tables S2-3. Examples of exposures and phenotypes are displayed in Tables S4-5.

**Figure 1.**
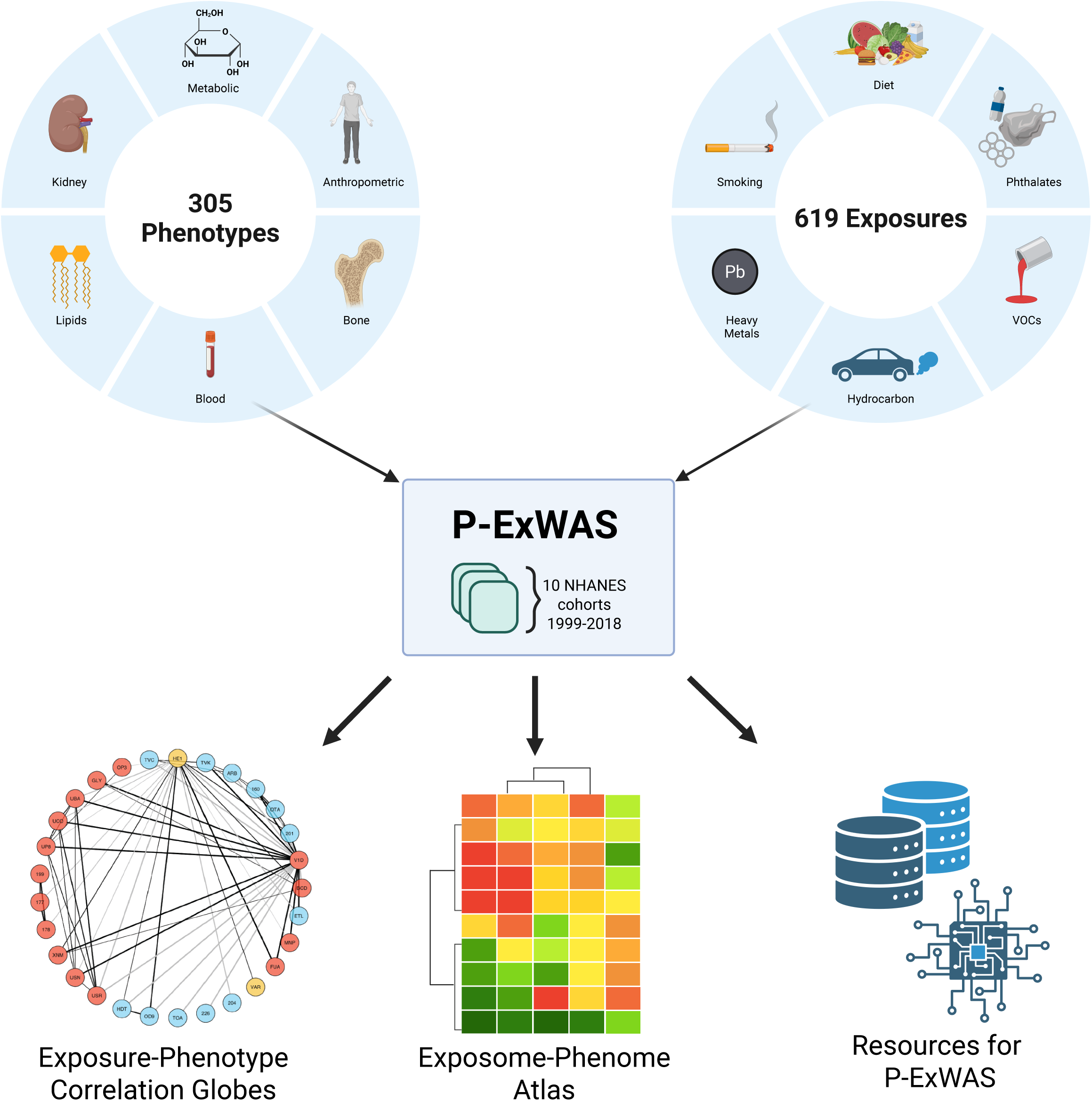
Schematic of conducting systematic Phenotype-by-Exposome Wide Association Studies (P-ExWAS). Top left panel describes samples of the phenotypic domain comprising 305 phenotypes. Top right panel describes samples of the exposomic domain comprising 619 exposures. These data are harmonized across 8 cohort samples of the *National Health and Nutrition Examination Survey* 1999-2018. Bottom panel describes resources to describe the architecture of phenome-exposome associations, including exposome globes, the Exposome-Phenome Atlas, and digital resources for conducting P-ExWAS (database and software).

### Exposure-wide associations across the phenome

We conducted an “phenotype-by-exposome-wide association study”, whereby each exposure is related with each phenotype ^15^. We used survey-weighted regression to associate phenotypes with all exposures under 9 different modeling scenarios that adjust for demographic and social attributes: the (1) main reported model, which consists of age, age^2^, sex, income (household income index divided by the poverty level), ethnicity (5 groups) and education (3 groups: above high school, high school equivalent, and below high school), and survey year (e.g, 1999-2000, 2001-02, 2003-04, 2005-06, 2007-08, 2009-10, 2010-11, 2012-13, 2014-15, 2016-17 as a categorical) (2) base model, with no adjustments, (3) sex and survey year, (4) age and age^2^ and survey year (5) sex, age, and age^2^, and survey year (6) ethnicity and survey year, (7) income, education, and survey year (8) age, age^2^, sex, and ethnicity and survey year, and (9) age, age^2^, sex, income, education, and survey year.

We scaled all continuous exposures and phenotypes by their SD (see Methods) and ran regression models to obtain standardized β-coefficients, p-values, and R² (Figures 2-4) Categorical exposures were compared to predefined reference groups. Significance was defined by a Bonferroni threshold (α≈4×10⁻L) and the Benjamini–Yekutieli FDR.

**Figure 2.**
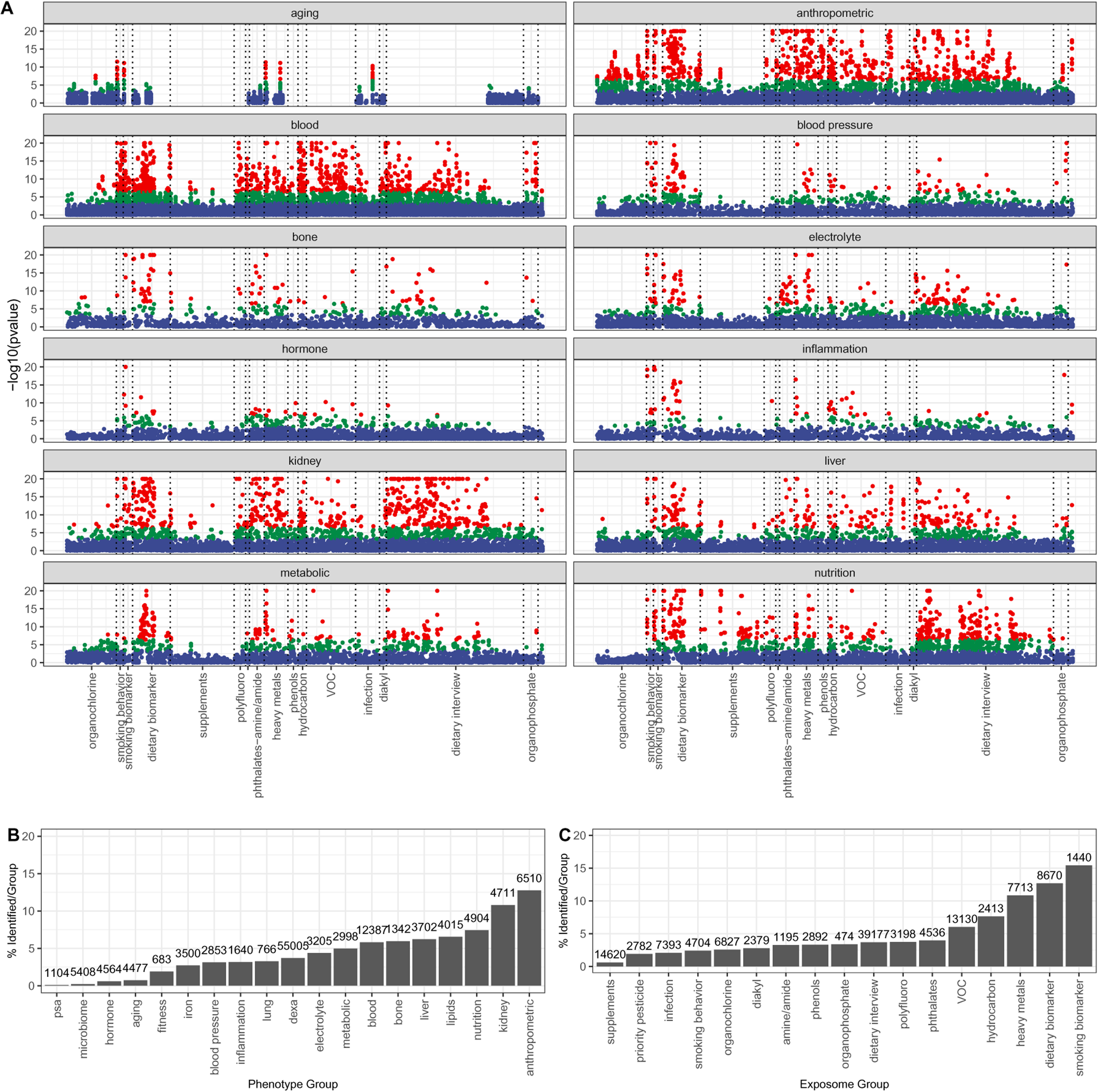
Associational architecture of the exposome on the phenome. A.) Upper left panel shows the –2-sided log10(p-value) vs. the exposure type. P-values are not shown corrected for multiple hypotheses. B.) number of significant phenotype-exposure associations per phenotype category (total E associations shown in text above bar), C.) number of significant phenotype-exposure associations per exposome category (total phenotype associations shown in text). Red: associations below Bonferroni (4e-7), Green: associations below FDR (5e-4) but greater than Bonferroni, Blue: associations greater than FDR.

**Figure 3.**
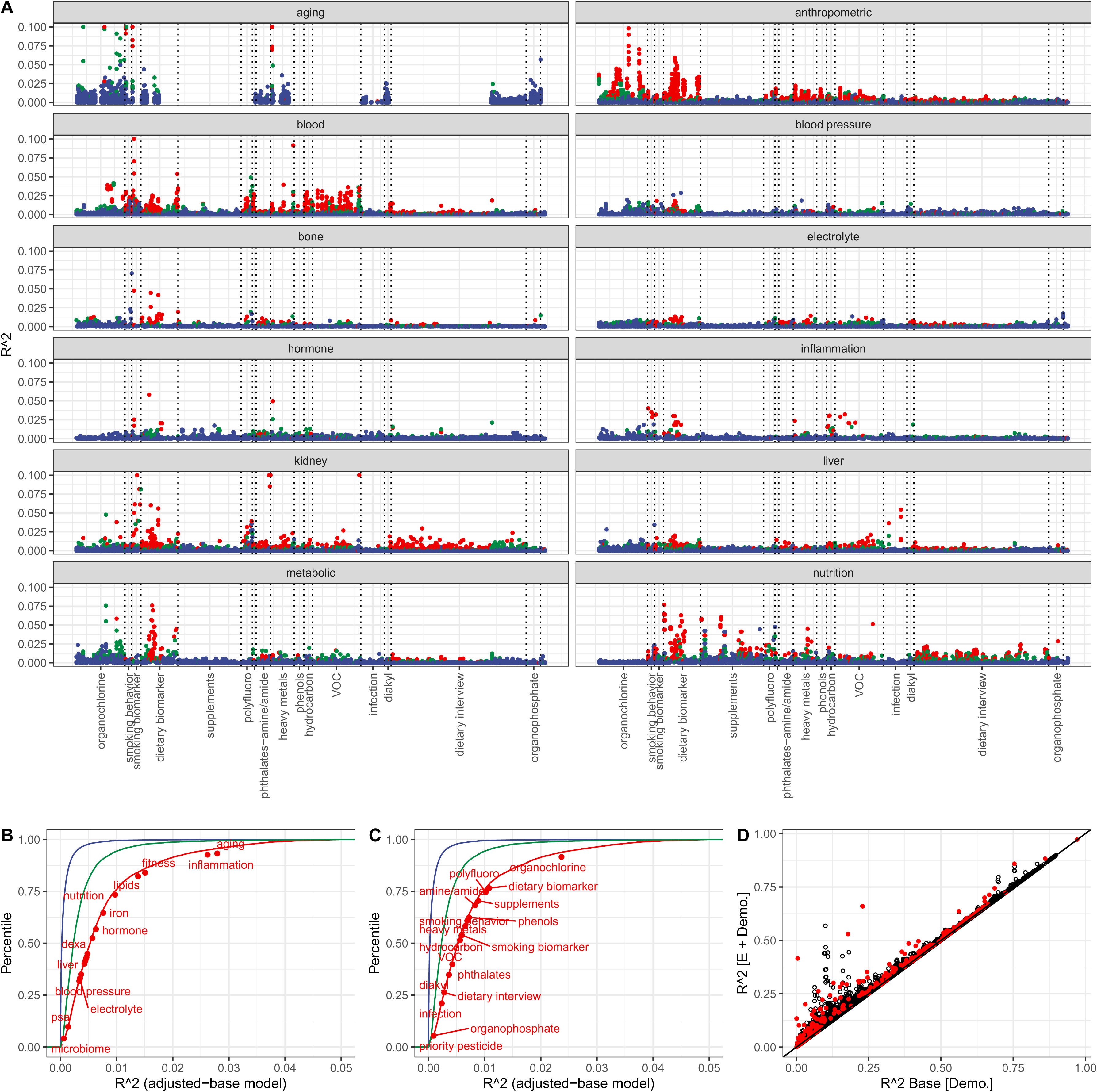
Variance explained by the exposome across phenotype groups. A.) R-squared (R^2^) for exposure across exposure categories and phenome categories. B.) Cumulative distribution of R^2^. The median R^2^ for each phenotypic category is annotated. C.) Cumulative distribution of R^2^ across 119k phenotype-exposure associations. The median R^2^ for each exposome category is annotated. Colors for A-C: Red: associations below Bonferroni (4×10^−7^), Green: associations below FDR (5×10^−4^) but greater than Bonferroni, Blue: associations greater than FDR. D.) R^2^ attributable to the exposure versus only to Demographics (age, age^2^, ethnicity, income, education, and sex). Red: R^2^ attributable to multiple simultaneous exposures (up to 10). Demo.: Demographics (age, age^2^, ethnicity, income, education, and sex)

**Figure 4.**
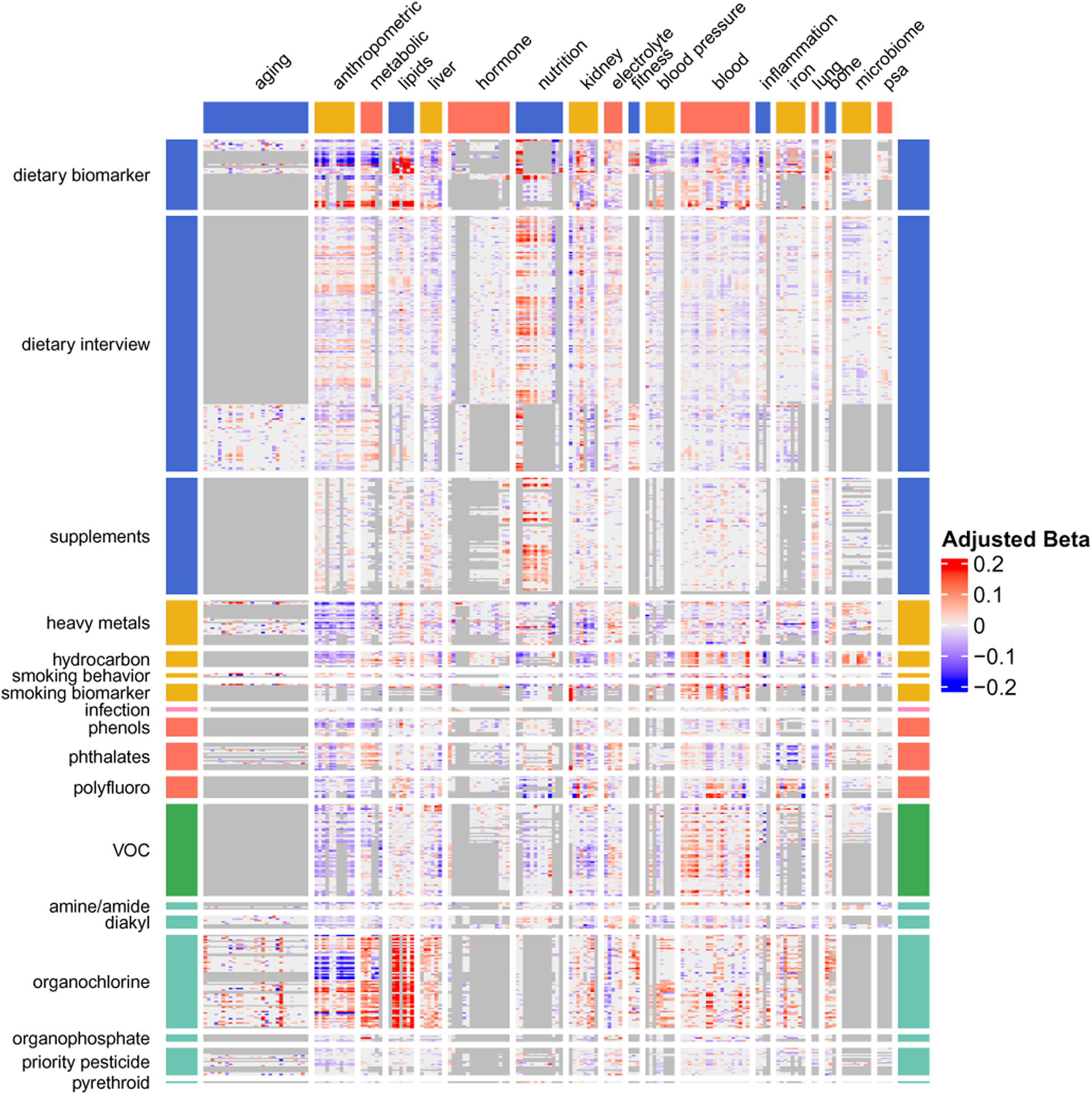
Phenome-Exposome Atlas. 305 phenotypes across 18 categories are depicted column wise, 625 exposures across 18 categories are depicted row wise. Each entry in the matrix is the linear association relationship (“Adjusted Beta”) between exposure and phenotype. Grey shading denotes association that could not be estimated due to pair-wise missingness or total sample size less than 500.

The number of associations across all phenotype-exposure associations that passed the Bonferroni threshold was 5,674 (5% of 123,774) (Figure 2A, blue shaded points, Figure S2). The p-value corresponding to a Benjamini-Yeukuteli correction of 5% was 5.1×10^−4^ (Figure 2A, blue and red shaded points). The total number of associations that exceeded the FDR of 5% was 15,386 (12%).

The total number of tests per phenotype (n=305) conducted was 16 to 654 (median of 397). The average percent of associations (per phenotype) that were Bonferroni significant was 5% (range of 0.25% to 20%). The most associations were found for serum bilirubin, waist circumference, and body mass index (20% of tests for these phenotypes were significant for over 640 total tests).

We observed a large range of identified associations by exposome or phenome category (Figure 2BC). For example, the anthropometric phenome category saw the highest number of associations (13% of phenotypes in this category had a Bonferroni significant association)(Figure 2B, Figure S2). Of exposome variables, smoking and dietary/nutrient biomarkers were implicated in the most phenotype-exposure associations, ∼15% and 13% respectively (Figure 2C, Figure S2).

### Phenotype-exposure associations replicate across cohorts

For reporting of main results, we combined cohort samplings across survey waves to maximize power. We also can examine associations within each of the survey samples to estimate the approximate “replication rate”. We estimated the rate of “replication” or the number of times an association appeared to be nominally significant at a pvalue threshold of 0.05 in greater than 1 survey sampling, which we call a “replication rate”. Of the 5,674 associations that were Bonferroni significant, the replication rate was 41% (n=2,321). In contrast, for those that did not achieve FDR nor Bonferroni significance, the replication rate was 0.8% (n=867).

Across the atlas of associations, we found that an association (at a p-value threshold of 0.05) occurred in two surveys 5% of the time. To contrast, if a phenotype-exposure association achieved FDR significance across all surveys (Figure 2) p-value significance was achieved in greater than 2 surveys at least 20% of the time (Extended Data Figure 2). However, phenotype-exposure replicated rates vary depending on the number of surveys available for a phenotype-exposure association. Specifically, for FDR-significant phenotype-exposure associations assessed in only 2 surveys were found in both surveys 39% of the time at p-value less than 0.05 (Extended Data Figure 2). For associations that were at least FDR significant, the median I^2^ was 0, 5, 0, 26, 6, 14, 14, 20, and 18% for associations in 2, 3, 4, 5, 6, 7, 8, 9, 10 surveys. We also assessed the percent that were nominally significant in multiple survey waves. For the 1,211 associations estimated in 10 survey waves (e.g., they were tested in each of the 10 surveys), there were 76%, 11%, 4%, 2.5%, 1%, 1%, 1%, 0.5%, 1%, 1% of associations that were nominally significant in exactly 0, 1, 2, 3, 4, 5, 6, 7, 8, 9, or 10 surveys. In other words, the replication rate among 1,211 associations estimated in 10 different cohort samples was 13% (n=161/1211).

See Supplementary Results for heterogeneity of association across survey samples (Figure S3).

### Variance explained of the exposome

We estimated the variance explained (R^2^) attributable to the exposure variable (after subtracting the potential role of demographic attributes [Methods]) (Figure 3B-D). Demographic factors, including age, age^2^, ethnicity, income, education, and sex explained a large range of overall phenotypic variation, from ∼0 to 80% (Figure 3D, x-axis). In comparison, single exposures added a median of 0.14% (Figure 3A, D).

The median R^2^ for all associations that were Bonferroni significant was 0.6% (IQR: 0.3 and 1%; 5th to 95th percentile 0.1% to 3.6%) (Figure 3BC). The median R^2^ was 0.02% for non-significant associations. We observed a range of R^2^ by exposure associations across domains of the phenome and exposome (Figure 3BC). For example, phenotypes in the inflammation category had exposures that explained 3% of variance on average across all phenotype-exposure associations (Figure 3B) that were Bonferroni significant. For exposures, pollutant factors explained on average 0-3% of variation across all phenotypes. Organochlorine exposures explained ∼3% of variance on average across all phenotypes. On average, dietary biomarkers accounted on average 1% of variation; however, dietary factors measured through an interview explained on average 0.5% of variation in phenotype.

Second, we also estimated the R^2^ due to the additive contribution of 20 exposures simultaneously. For phenotypes that had greater than 20 exposures associated at FDR-level of significance, we imputed exposure data where missing (see Methods). When considering 20 exposome factors simultaneously in 119 phenotypes, the median variance explained is 3.5% (interquartile range 1.8% to 7.9%), greater than the median R^2^ for single exposures (Table S6). When considering all phenotypes with less than or equal to 20 exposures in the model, the median R^2^ was 1.6% (interquartile range 0.7 and 3.5%) (Figure 3D, red points).

The maximum multiple exposure R^2^ estimated was 43% for triglycerides (Figure 3D, red points). Triglycerides are an important clinical phenotype used to screen for cardiovascular disease. We found that aggregate exposome, particularly lipophilic dietary and pollutant-related exposures, described large variance in levels of triglycerides in the US (R^2^ of 43%, the largest of any phenotype) (Table S6-7), even after adjusting for total cholesterol. Of the 20 variables in the model (Table S8), a trans fatty acid (Trans,trans-9,12-Octadecadienoic acid), alpha-tocopherol, and gamma-tocopherol independently contributed the most to the variance and were positively associated with triglycerides (Table S8).

### An atlas across the exposome and phenome

Association sizes correspond to the amount of the change in 1 standard deviation (SD) of exposure with a change for a 1SD change for the log-transformed exposures (for continuous exposures) or versus the reference group (for categorical variables) (“adjusted beta”, Figure 4). For associations between Bonferroni significant exposome factors and phenotypes (association sizes for a 1SD change in exposome factor), the 5th to 95th percentile range was –0.17 to 0.19 (0.03 to 0.24 in absolute values).

### Dense correlational web of the exposome

Exposures exhibit a dense correlation web (Figure 5AB). The median correlation between exposures was 0.01 and the median absolute value correlation was 0.05 across all correlations. For exposure-exposure correlations that passed the Bonferroni threshold (p < 0.05/201,265), the median for Bonferroni-corrected correlations (alpha threshold of 2e-7) was 0.19 and the median absolute value correlation was 0.21 (Figure 5C); the interquartile range of Bonferroni-corrected correlations was 0.08 to 0.37 (Figure 5C) (0.11 to 0.38 for the absolute values). The 95th percentile reached a correlation of 0.69 (0.69 for the absolute values).

**Figure 5.**
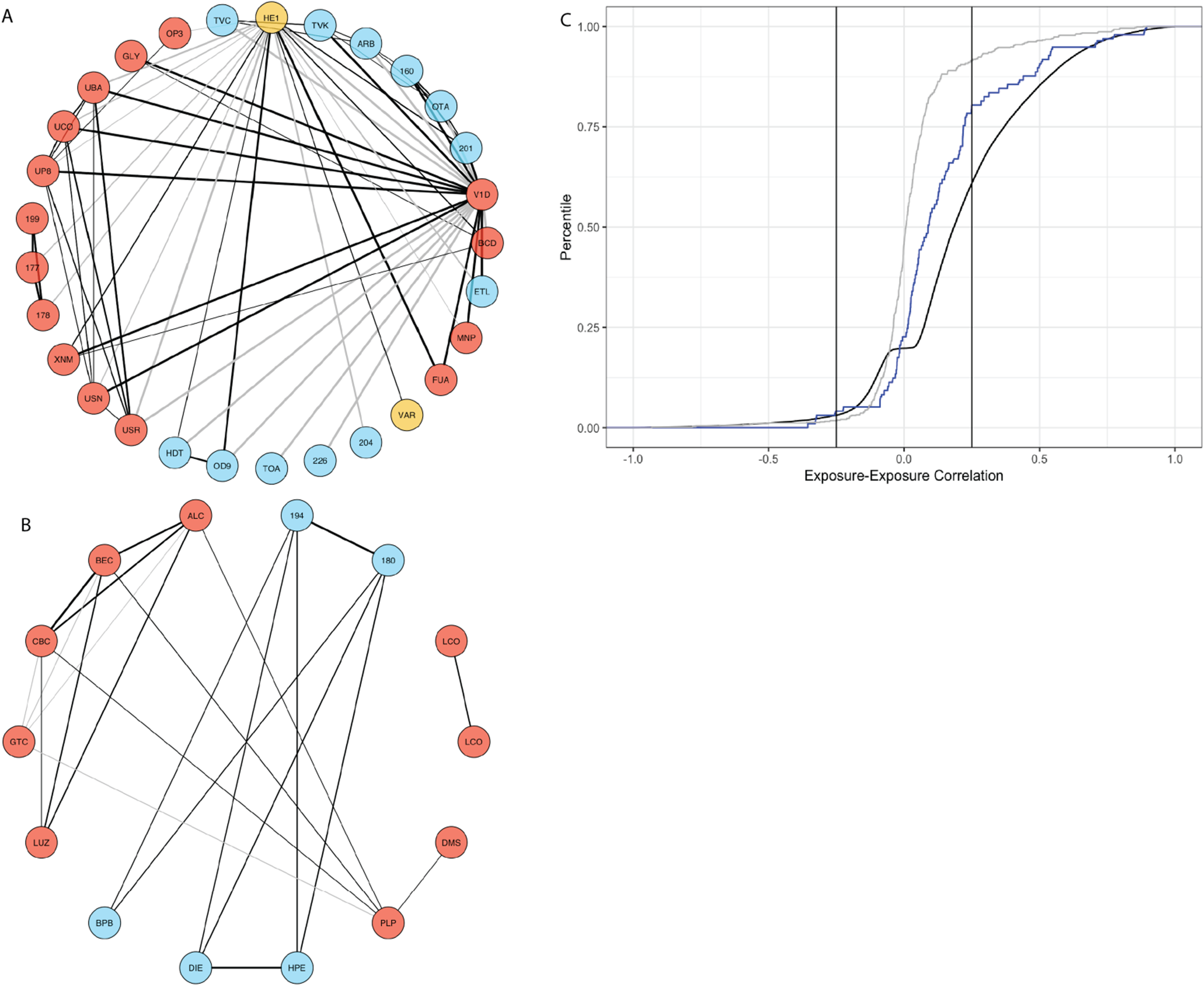
Exposome-exposome correlational globe and distribution of exposure-exposure correlations across the exposome. A.) An example Exposome Globe depicts exposure factors and their correlations chosen randomly (thresholded for absolute values of correlations above 0.25). B.) Exposome Correlation Globe for exposures associated with Hemoglobin A1C or BMI. Node colors include: pollutants (red), infection (yellow), nutrients (red). C.) Distribution of exposure-exposure correlations. Grey color denotes randomly selected correlations. Blue line denotes exposome correlations for exposures associated with BMI or Hemoglobin A1C. Black denotes correlations that achieved Bonferroni significance. Lines depict correlations at –0.25 and 0.25.

The dense correlational web is described for a sampling of exposures in a correlation globe (Figure 5AB, see methods). Figure 5A depicts 50 randomly selected correlations sampled from all 201,265 correlations. Figure 5B depicts exposures identified in their association with BMI and Hemoglobin A1C%. Correlations are only drawn between nodes (or exposures) whose absolute value of correlation was greater than 0.25; these correlations are among the top 15% of the distribution of correlations (Figure 5C). Correlations for exposures associated with BMI and A1C% overall have larger correlations than a randomly selected subset (Figure 5C, blue).

### Demographic adjustment influences association sizes

Next, we computed the difference between adjusted associations (seen in Figure 4) subtracted from univariate estimates to assess the impact of adjustment. The average difference between a minimally adjusted and a fully adjusted model and each scenario was 0.01 (Extended Data Figure 3A), demonstrating some bias due to demographic adjustment. The standard deviation sizes were largest for the fully adjusted minus univariate model (SD: 0.1), fully adjusted minus ethnicity model (SD: 0.1). The SD were least for the fully adjusted minus the age, sex, and ethnicity models and the fully adjusted minus the age, sex, and income/education models (Extended Data Figure 3A). See the Supplementary results for findings regarding specific exposures.

For some associations, the sign of association was opposite depending on the demographic correction scenario. Of the significant findings, 932 out of 5194 (15% of total significant Bonferroni identified pairs) exhibited a switch of coefficient sign between the univariate model (a model with no demographic or social factor adjustment) and the adjusted model. For example, BMI and blood cadmium, had positive associations (e.g,, for an increase in exposure, there is linear increase in BMI); however, when controlling or adjusting for factors in the “main” model, the association becomes stronger (e.g., the standard errors are reduced) and opposite in direction (Extended Data Figure 3B). The difference between adjusted associations also differed per exposome domain (Figure S4).

### Consistency of associations across exposure categories

Self-reported dietary nutrients and variables dominate nutrient exposure assessment in epidemiological studies. Overall, we found that 1,452 phenotypes had Bonferroni significant associations with self-reported questionnaire derived nutrient variables; however, among these phenotypes, they exhibited a median R^2^ of only 0.2%.

Next, we hypothesized that a dilution of correlation size was due to measurement noise. Self-reported dietary nutrients were assessed on two days. The median correlation across 69 dietary nutrients recall in day 1 vs day 2 was 0.36 (IQR range of 0.28 and 0.43). We estimated the correlations of associations (e.g., beta-carotene-P association on day 1 vs. beta-carotene-P association on day 2) (Extended Data Figure 4A). Across all levels of significance, we observed a 0.84 correlation between day 1 self-report vs. day 2 (Extended Data Figure 4A).

Dietary biomarkers, on the other hand, had a larger median R^2^ of 1% across the 1101 phenotypes with Bonferroni significance, 5 times larger than their self-reported counterparts. We also estimated the correlation between biomarkers and their self-reported counterparts (e.g., day1/day2 average of trans beta carotene vs. serum trans beta carotene) (Extended Data Figure 4B). The correlations between self-reported and biomarkers were smaller, and we observed a Pearson correlation of 0.52. For those blood nutrient variables that were Bonferroni significant, we observed a correlation of 0.60.

Blood and urine pollutant biomarkers reflect the biological relationship between exposure and excretion. We observed a positive and strong correlation between associations. For example, for blood vs. urinary biomarkers, we observed a 0.72 Pearson correlation. When only considering blood biomarkers that were Bonferroni significant, the phenotype-exposure associations between blood vs. urine biomarkers was 0.78 for cadmium, 0.96 for cotinine, and 0.71 for mercury (Extended Data Figure 4C).

### Consistency of exposome associations for lung function

Smoking is a strong risk factor for reduction of lung function, such as the amount of air expelled (e.g., *FEV1*). Smoking-related biomarkers such as NNAL and serum cotinine indeed show negative associations with FEV1, consistent with prior evidence linking tobacco exposure to reduced lung function. Urinary NNAL (4-(methylnitrosamino)-1-(3-pyridyl)-1-butanol (NNAL) is a tobacco-specific nitrosamine) showed a stronger negative association with lung function (Forced Expiratory Volume, 1 second, FEV1 (–0.06 per SD, R² = 0.2%) compared with serum cotinine (–0.03 per SD, R² = 0.08%). The difference in association between cotinine and NNL is consistent with their biological properties: cotinine, a short half-life metabolite of nicotine, primarily captures recent exposure and is subject to greater day-to-day variability, whereas NNAL, a metabolite of the tobacco-specific nitrosamine with a half-life of 10–16 days, provides a more stable marker of cumulative exposure. Nevertheless, numerous densely correlated exposures emerged associated with FEV1 (Extended Data Figure 5).

### Exposome correlates of methylation and cognitive aging

We deployed our ExWAS procedure across timely biomarkers of aging, including epigenetic age (e.g., Horvath’s biological age predictor and GrimAge [which includes smoking status]^17,18^). We also interrogated the indicators of cognitive recall (digit substitution test), as they are used in the clinic to stage cognitive decline in the elderly (Figure S5). Volatile organic compounds, smoking indicators (cotinine), and physical activity had the strongest associations with cognitive function (Figure S5AB). We found shared exposure associations (or, “shared architecture”) between better cognitive function and other phenotypes interrogated in the population, including higher exhaled nitrous oxide (shared correlation of 0.35) and with urinary creatinine (Figure S5C).

The strongest signals for accelerated epigenetic age (such as *GrimAgeMort*) was associated with smoking, heavy metals, and physical activity behavior. Among these domains physical activity explained the most variance (less than 1% in R^2^); however, in terms of aggregate total exposomic risk, 10 exposures explained 10% of the variance in GrimAgeMort (Table S6).

### Age by exposome interactions

Recognizing that the effect of an exposure may vary depending on an individual’s age at the time of exposure, we modified the *ExWAS* procedure to consider age by exposure interactions (e.g., does the association size differ for individuals at different ages?, see Methods). In summary, the inclusion of exposure-by-age interactions resulted in only marginal improvements in the variance explained (R²) for most phenotypes (see Supplementary Results and Extended Data Figure 6A-C). Most of the additional variance for models that incorporate an interaction term are limited, although there are exceptions.

### Shared associational architecture between exposures

For a pair of exposures, shared “associational architecture” measures the correlation, or the similarity of their correlations across all phenotypes (Extended Data Figure 7AB). For example, the correlation between associations for blood trans– vs cis-beta-carotene was 0.98; in other words, the association coefficients across the phenome were very similar for those two exposures. The associational architecture between serum cotinine and 3-fluorene had 0.90 correlation.

Exposures from the same category (e.g., dietary interview, organochlorine, VOC, dietary biomarkers) tended to have similar phenotypic associations; the degree of shared associational architecture is larger within categories than across categories. For all correlations within exposure variables that were dietary biomarkers, the median absolute value of shared architecture was 0.2 (IQR 0.01 to 0.35). Similar shared associational architecture was observed within smoking biomarkers (0.2, IQR 0.08 to 0.35). The shared associational architecture between dietary biomarkers and self-reported dietary nutrients had a median absolute value correlation of 0.24. We examined the degree of shared associational architecture between phenotypes. The shared associational architecture between BMI and body weight was 0.98. BMI and cardiorespiratory fitness had opposite associational architecture (the sign of the correlations were opposite between BMI and fitness), a correlation of –0.83. Hemoglobin A1C% had an opposite architecture compared to HDL-Cholesterol (correlation of –0.54). See Supplementary Results for shared associations between blood and urine measured exposures.

### Comparison with genome wide association studies

We used data from participants of the UK Biobank to compare genetic versus exposomic predictions. We compared the variance explained due to ∼1M imputed genotypes from genome-wide association studies performed on 29 of the phenotypes^19^ examined here (Extended Data Figure 8; Table S7). Across the 29 phenotypes, the median incremental R² due to genetics was 7.9L% (IQRL2.8L% to 9.3L%; maximumL21L%) and the median incremental R^2^ due to exposome (20 exposome variables across 39 phenotypes) was 7.9% (IQR 3.1% to –12%; maximum 57%). We found that the multiple exposome factors, when modeled simultaneously, had explained variance comparable to the entire genomic array across the phenotypes (Extended Data Figure 8). Specifically, 55% (n=16) phenotypes had higher exposomic versus genetic R^2^ (Extended Data Figure 8). For example, the total genetic (1M common SNPs) and exposomic (20 factors) variance explained for BMI was similar, ∼10% for both.

We benchmarked the atlas against three exposure-wide analyses, and found our findings were concordant (directionally consistent and robust pvalues) with prior published data found in refs ^20^ ^21^ ^22^.

## Discussion

Our systematic mapping of the exposome onto the phenome reveals three insights with direct clinical and biomedical implications. First, robust environmental signals are highly concentrated in cardiometabolic and pulmonary phenotypes used to stage and gauge care, establishing lipids (specifically triglycerides), glycemic markers, and lung function (FEV1) as the highest-yield targets for our data-driven environmental risk assessment. Objective nutrient biomarkers and lipophilic pollutants are reproducible correlates of body mass index, glycated hemoglobin, and lipid profiles. Triglycerides stood out as the phenotype most strongly linked to multi-domain exposure patterns, with trans fatty acids, banned persistent pollutants (e.g., polychlorinated compounds), and vitamin E isoforms among the most informative contributors, suggesting that lipid risk assessment – important for staging cardiovascular disease – may be sensitive to integrated dietary and pollutant chemical contexts. In pulmonary traits, tobacco-specific biomarkers showed stronger and more stable associations with reduced lung function than short-lived nicotine metabolites, supporting the clinical utility of longer half-life biomarkers when refining smoking-related risk for forced expiratory volume in 1 second (FEV1) and related outcomes (e.g., chronic pulmonary disorder). Importantly, we also demonstrate that while single exposures have modest association sizes, aggregate “poly-exposomic” profiles explain phenotypic variance comparable to genome-wide polygenic scores; this suggests that multi-factor environmental exposome integration is required to meaningfully improve precision risk models beyond age and sex. Third, we identify a critical reliance on objective measurement: biomarkers (e.g., serum nutrients, urinary tobacco metabolites) consistently revealed biomedical associations that self-reported history failed to capture. Collectively, these findings move the field beyond fragmented and non-objective associations, defining the specific clinical domains and measurement modalities necessary to operationalize the exposome in biomedical research to evaluate medical decision-making.

Our findings have several implications. Estimating association sizes and their replication rate across exposure domains helps prioritize which exposures are most likely to yield clinically meaningful signals and can guide study design and power planning for ExWAS in new cohorts (Table 1). Most of the exposome tabulated here adds little incremental clinically-relevant predictive value for many phenotypes, but a smaller set especially in cardiometabolic and smoking-related domains appear more promising for refining risk equations. We note that cancer-related phenotypes are underrepresented in our atlas and are ripe for further research.

Our data suggests that clinically useful environmental risk stratification will more often require integrated, multi-exposure models (e.g., ^23^)) rather than isolated markers. Demographics remain essential for risk adjustment, and modest exposome signal strength likely reflects measurement limitations and the cross-sectional nature of many exposure assessments.

Most exposures show broad, non-specific associations across phenotypes and are often correlated with other exposures ^24–26^, complicating causal interpretation and attribution, underscoring the need to view high-priority signals in the context of exposure “mixtures”, globes, and bundles. This is exemplified by smoking, where biomarker indicators of the behavior with different half-lives may capture distinct time windows of exposure relevant to lung function; however, the exposures are all related to one another and make a complex globe.

Inferred exposure–phenotype relationships are sensitive to analytical choices and confounding control ^27^, especially for age, sex, and socioeconomic factors, reinforcing the need for transparent modeling and framing. Future work should test whether top signals persist under alternative adjustment strategies and in longitudinal settings, and the exposome field may need to configure model specifications per exposome or phenotype domain and explore mediation and interaction^28^.

Objective biomarkers appear more consistent and informative than self-reported measures, with strong concordance across blood and urine heavy metal indicators and far weaker signals for dietary recalls compared with nutrient biomarkers. This supports prioritizing standardized biospecimen assays when the goal is clinical translation for precision medicine or robust population surveillance.

We have several technical recommendations for implementation of *ExWAS* (Table S9). Exposomics, at present, is associational discovery. To move toward causal attribution, we recommend a triangulated strategy (e.g., ^29^) that prioritizes top exposure–phenotype pairs, ranked by replication rate, effect size, p-values, and vibration-of-effects ^30^, for targeted follow-up. Next, establish temporality in disease-specific longitudinal cohorts by measuring exposures at baseline and relating them to time-to-event or longitudinal trajectories. One can apply instrumental-variable approaches, including Mendelian randomization ^31^, where genetic variants serve as stand-ins for exposures. Third, aim for “functional exposomics” and measure more granularly and precisely, using proteomics, metabolomics, and methylomics to map exposure-responsive pathways and test mediation (e.g., ^32,33^). For behavioral exposure bundles (e.g., diet), investigators should move to randomized interventions.

Our study has limitations, with many directions to evaluate next. Despite cataloging hundreds of factors, we capture only a fraction of the total exposome; characterizing complex exposure–exposure and gene–environment interactions^34^ will require larger sample sizes and broader, high-resolution chemical profiling. While our reliance on objective biomarkers yields stronger signals than the self-reported or geospatial proxies often used in other biobanks, direct cross-cohort comparison is complicated by differences in sampling frames (e.g., volunteer bias in cohorts such as UK Biobank ^35^). Furthermore, our systematic evaluation suggests that many associations reported in prior candidate-exposure literature may be false positives ^9^. The cross-sectional design limits causal inference and the capture of cumulative lifetime exposures. Although we attempted to model participant age as modifying exposure-phenotype relationships, delineating critical windows of susceptibility or non-linear temporal dynamics ultimately requires longitudinal designs to distinguish chronic accumulation from acute reverse-causal effects. The exposomic architectures measured here are chronic, but they may also be acute, such as glucose response to diet and physical activity ^36,37^. Frontier studies will incorporate dynamic and personalized exposome and phenotype measurements such as continuous glucose monitoring devices to ascertain heterogeneity or person-specific responses to the exposome ^38^.

Our atlas will complement existing global efforts to document exposure phenotype relationships. For example, there are numerous biobanks/cohorts with phenotype measures, and to some extent, exposure measures such as those documented in the *Human Health and Exposure Analysis Resource* (HHEAR: https://hhearprogram.org/data-center) (Extended Data Table 1). A recent seruml⍰only exposome mapping in a Chinese cohort (You et al.) ^39^, prioritized breadth and population representativeness, interrogating 267 blood chemicals in 5,700 volunteers. We view our atlas as a complement to these efforts and will help to devise standards by which to catalog associations^40^ to enhance longer term reproducibility.

Emerging efforts are expanding the molecular exposome with increasingly precise high-resolution assays, including targeted panels and high-resolution mass spectrometry (e.g, ^41^), but the next major advance will be enhanced measurement paired with expanded study designs. In particular, repeated and personalized exposure profiling that spans chronic burdens and acute signals will be essential for moving to causal implication of the exposome in disease. Such measurement-rich longitudinal studies, will further prioritize the most promising exposure domains for follow-up, clarify temporality, explain personal heterogeneity, and identify modifiable drivers of clinical phenotypes. As these studies mature, the field will be positioned to define causally attributable exposures that can be targeted through behavioral, pharmaceutical, environmental, or policy interventions and/or incorporated into predictive models (e.g., ^42^) for individual risk assessment in the clinic. In this way, our *ExWAS* serves as a prerequisite for systematic, large-scale integration of exposomic information at the point of care.

## Acknowledgements

We thank Gary Miller for their review of early findings. We thank Dennis Bier. This study was funded through NIH grants, including NIEHS R01ES032470 (CJP, JPAI, AKM), NIDDK R01DK137993 (CJP, AKM), NIEHS U24ES036819 (CJP), USDA (CJP), and N000142412687 by the Office of Naval Research (JPAI). The funders had no role in study design, data collection and analysis, decision to publish or preparation of the manuscript.

## Author Contributions

CJP, AKM, and JPAI conceived of and wrote the initial draft of the paper. CJP obtained the data and analyzed the data.

## Tables

None.

## Online Methods

We systematically associated environmental exposures and phenotypes, called an Phenotype-by-Exposure Wide Association (P-ExWAS) ^15^, leveraging participant data from the Centers for Disease Control and Prevention (CDC) National Health and Nutrition Examination Survey (NHANES). Extended Data Figure 1 describes the pipeline workflow. To enhance replication, the findings are deployed as a database called a “Phenome-Exposome Atlas” (Table S1). Our study complies with the STROBE checklist.

We developed a R statistical package (“*nhanespewas*”) to conduct all analyses. The features of the *nhanespewas* includes (a) cataloging of phenotypes and exposures to associate within the NHANES surveys, (b) R package to associate all of the exposures with phenotypes using a survey-weighted linear model^43^ and providing a user-specified array of modeling assumptions (e.g., potential confounder and covariate choice), (c) aggregating pairwise associations across independent surveys (a meta-analysis) to output an overall estimate across surveys and assess replicability, and (d) providing a browsable database (*Phenome-Exposome Atlas*) of summary statistics that contain the overall association size or correlation across all survey years interrogated, the standard error of the association, p-value, and variance explained attributable to the exposure (and after consideration of potential adjusting covariates). The code and package is located here: https://github.com/chiragjp/nhanespewas. An introduction to the package is found here: pe_quickstart.Rmd (https://github.com/chiragjp/nhanespewas/blob/main/pe_quickstart.Rmd)

### Statistics and Reproducibility

This study utilized an observational, cross-sectional design leveraging 10 serial waves of the National Health and Nutrition Examination Survey (NHANES) from 1999–2018. No statistical method was used to predetermine sample size. Sample sizes were determined by the availability of participant data within the public-use NHANES database. To ensure robust associational estimates, we limited our analysis to phenotype-exposure pairs with a minimum of 500 participants across at least two survey cycles. Post-hoc power calculations were performed to determine the minimum detectable effect sizes (R^2) for the resulting sample ranges (median N = 7,464), as detailed below.

No data were excluded from the analyses, except for the pre-specified filtering criteria: phenotype-exposure pairs were required to have at least 500 participants and exist in more than one survey wave to be included in the atlas. Participants with missing values for required demographic covariates in a specific model were excluded from that specific regression (complete-case analysis).

As this was an observational study using secondary public health data, the experiments were not randomized and the Investigators were not blinded to allocation during experiments and outcome assessment.

Statistical analyses were conducted using survey-weighted linear regression to account for the complex, multi-stage sampling design of NHANES. Standardized coefficients, p-values, and R^2 were calculated for each pair. Multiple testing was accounted for using both the Bonferroni family-wise error rate (∼ 4×10^-7) and the Benjamini–Yekutieli False Discovery Rate (FDR < 5%). Missing data for multi-exposure models were handled via Multiple Imputation with Chained Equations (MICE). All analyses were performed using R (version 4.x) and the survey package.

To ensure the reproducibility of these findings, the full analytic pipeline is provided as an open-source R package, nhanespewas, available at [GitHub link]. All summary statistics, including the Phenome-Exposome Atlas, are available in a searchable digital database and archived via Figshare.

#### Attaining and cataloging participant data from the National Health and Nutrition Examination Survey

First, we download all participant data and data dictionaries, encompassing 10 surveys (approximately 10,000 total participants per survey) (Extended Data Figure 1A). Participant data in NHANES are divided into different tables in 5 components (demographics, diet, laboratory, questionnaire, and examination). We next categorized each variable downloaded into being a “phenotype” or an “exposure”. We defined exposures as exposures if they are (a) exogenous in origin (e.g., pollutants) (b) biomarkers of exposure (e.g., cotinine),(c) reflective of lifestyle or behavior (e.g., self-reported food intake). While many of these variables do exhibit heritability (e.g., have genome-environment correlation), it is secondary as the origin of the factor is still external.

We processed some phenotypic variables and exposure variables. Blood pressure was a phenotypic variable whose measurement was repeated multiple times: we took the average of the measurements. Physical activity questionnaire information was collected in a series of questionnaire items; we processed these measurements to be in total metabolic equivalent hours (see websites: ^44–46^). We processed multiple smoking variables (see ref. 26) to be a variable that denotes past, current, or never smoking. We next tabulated the sample size for each exposure and phenotype variable pair, used in downstream pipelines.

We programmatically ingest NHANES data dictionaries and public data files for 10 cycles (1999–2018) into a SQLite database and build a canonical catalog that maps each exposure (E) and phenotype (P) to all cycle-specific variable names and labels referring to the same construct (as above). Code paths: download/ (data & dictionary retrieval) and select/ (catalog building, sample-size tables). Tutorials (pe_quickstart.Rmd, pe_catalog.Rmd) illustrate end-to-end usage. The repository and compiled database links are provided in Data/Code Availability. When NHANES supplies both conventional and SI variables (e.g., LBD*SI), we convert to a common unit before transformation; for categorical variables whose levels changed across cycles (e.g., education), we recode to harmonized bins listed in the catalog (Methods and Table S2–3). For analytes with known method revisions, we preferentially use NHANES-standardized variables when available—for example, LC-MS/MS-standardized 25-hydroxy-vitamin D (LBXVIDMS/LBDVIDMS; bridging equations applied by CDC) and IDMS-standardized serum creatinine—so that pre– and post-change data are comparable on a common scale. For laboratory measures, NHANES provides per-analyte LLOD and an “…LC” comment code. Values below LLOD are set to LLOD/√2, and the proportion <LLOD is tracked per analyte and cycle to aid interpretation and sensitivity analyses. Day-1 and day-2 dietary recalls are treated as distinct exposures; we present concordance analyses of P–E coefficients across days and (where available) compare interview-based nutrients with their biomarker counterpart. This avoids conflating changes in the underlying USDA nutrient databases with exposure measurement error. Vitamin D: we use CDC’s LC-MS/MS-standardized variables (LBXVIDMS/LBDVIDMS) across waves to harmonize measurements from earlier RIA methods to LC-MS/MS equivalence. Creatinine: we use NHANES IDMS-standardized serum creatinine (and NHANES-recommended corrections for older cycles where applicable). These steps follow CDC/NCHS analytic notes.

To ease interpretation, we categorized each cataloged phenotype or exposure variable into different categories. The categories of phenotypic variables (n=305) that included anthropometric (n=11, e.g., height), aging (n=29, e.g., telomere length and epigenetic aging), blood parameters (n=19, e.g., basophils number), bone (n=3, total calcium), dexa (n=136, e.g. lean fat), electrolyte (n=5, e.g., bicarbonate), fitness (n=3, e.g., Vo2max), hormone (n=17, e.g., iodine), inflammation (n=4, e.g., C-reactive protein), iron (n=8, e.g., iron), kidney (n=8, e.g., urinary creatinine), lipids (n=7, e.g., Apolipoprotein B), metabolic (n=6, e.g., glucose), microbiome (n=16, e.g., observed OTU mean), nutritional status (n=13, e.g., folate), and PSA (n=4, e.g., prostate specific antigen ratio).

The categories of exposome variables (total n= 619) included volatile organic compounds (VOCs, n=64, e.g., nitromethane), amine/amide (n=5, e.g., acrylamide), diakyl (n=9, e.g., dimethylphosphate), dietary biomarkers (n=49, e.g., vitamin B12), nutrients from dietary interview (n=178, e.g., alpha-carotene), heavy metals (e.g., blood lead), hydrocarbons (e.g., 1-napthol), infection (n=31, e.g., urinary chlamydia), organochlorine (n=65, e.g., PCB199), organophosphate (n=5, e.g., Acephate), phenols (n=13, e.g., e.g. urinary tert phenol), phthalates (n=19, e.g., mono(carboxynoyl) phthalate), polyfluorinated compounds (n=15, e.g., 2-(N-Methyl-perfluorooctane sulfonamido) acetic acid), priority pesticides (n=19, e.g., 2,4,5-trichlorophenol (ug/L)), pyrethoid (n=1, e.g., oxypyrimidine), smoking behavior (n=9, e.g., are you a current, ever, or never smoker?), smoking biomarker (n=12, e.g., cotinine), supplements (n=83, e.g., number of supplements reported). the entire list is in Table S2-3.

#### Model assumptions and data processing

The core functions of the package we now describe are available in the *quantpe.R* and *petable.R* (Extended Data Figure 1A). We model the relationship between an exposure and phenotype pair in the population with the following linear model:

Pp = intercept + bi*Ei + error, where Ei is an individual environmental factor or exposure of the exposome, the intercept is the average value of the phenotype when the exposure value is 0, and the b term denotes the association between the exposure and the phenotype. The lowercase p and i indexes a specific phenotype exposure respectively. We assume the error is also normally distributed with zero mean and standard deviation of 1. We modify this core model in a few ways to consider multiple adjustment scenarios and transformation of the Pp and Ei variables.

We transform the exposure and phenotype variables to ensure the comparability of estimates (e.g., bi) across all pairwise phenotype-exposure associations (Extended Data Figure 1B). Our pipeline inputs phenotypes, such as weight, body mass index, glucose, are quantitative and continuous valued variables. All phenotype variables are “scaled” (mean subtracted and divided by the survey-weighted standard deviation). Our pipeline also allows for rank based inverse normal transform, commonly used in genome-wide association studies ^47^.

We transform the exposures (*Ei*) in the following ways:

(a) for continuous and blood or urine biomarkers of exposure, we log transform (base 10) and scale (mean subtracted and divided by the survey-weighted weighted standard deviation). To account for zero values before log10 transformation, we add a small constant equal to the smallest non-zero value.
(b) for categorical exposure variables, we chose the largest category as the reference group, and analyzed the categorical variable in the same model
(c) for ordinal variables, we kept as ordinals and analyzed as a numbered rank (e.g., 1-3).

#### A pipeline to conduct survey-weighted associations

The NHANES is a complex and multi-stage cross-sectional survey designed by the National Center for Health Statistics; the multi-stage survey design provides US-wide generalizability for phenotype-exposure associations. Because of the complex study design, unique analytics approaches must be employed to ensure accurate association mapping between exposure and phenotype (versus approaches that assume a simple random sample). NHANES is sampled hierarchically: the first stage of sampling consists of regions of the country, or primary sampling unit (PSU) that are selected proportional to the average age, sex group prevalence, race and income. The second stage of sampling includes segments and households. Segments within the PSUs that are consistent with census blocks. The third stage of sampling includes individual participants. Each participant is also a weighted sample, such that it is proportional to the number of people in the population that that person represents by age, race, income, and non-response rate.

Our pipeline iterated through all combinations of phenotypes and exposures in the NHANES:

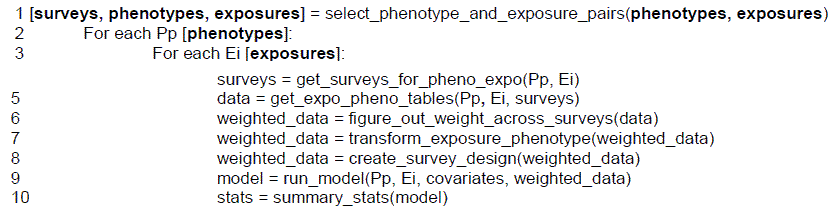

The NHANES participant data is organized into multiple tables based on properties of the measures that include the type of measure (e.g., dietary indicator or biomarker of exposure) on a subsample of the overall sample (Extended Data Figure 1A). Specifically, demographic variables, such as age, sex, race/ethnicity, and income are measured on the overall and entire sample; however, some assays of the exposome and phenome are measured on yet a subsample. A tutorial on the ExWAS approach in R is seen here: https://github.com/chiragjp/nhanespewas/blob/main/exwas_tour.Rmd

First, in line 1, we select pairs of phenotypes and exposures that have at least 500 measured participants. We further only select pairs that are present in more than one survey sampling. In lines 5-10 (Extended Data Figure 1B-D), we developed functions that query and merge the three or more data tables that contain the (1) phenotype and (2) exposure variables and (3) demographic variables and calculate the subsample weights to create a new NHANES-specific data table structure (see functions: *get_expo_pheno_tables*, *figure_out_weight* in the package). Next, we needed to combine data across multiple survey samplings and compute the new sample weights. For each survey, we calculated the subsample weights are selected by the smallest subsample being analyzed. For example, to estimate an association between fasting glucose (P) and urinary phthalate (E) biomarker, the data table that contains the smallest sample size will be the sample weight chosen. Last, we averaged the survey weights in accordance with the guidance provided by National Center for Health Statistics ^48^.

NHANES public-use data do not include state/county geocodes. All analyses used the survey R package with the provided PSU and strata identifiers and the appropriate exam/subsample weights for each exposure–phenotype pair. This approach yields design-consistent estimates that are generalizable to the U.S. non-institutionalized population and absorbs regional composition via post-stratification.

Before finalizing covariates, we examined the age–phenotype relationship using nonparametric smooths on representative phenotypes across categories (anthropometry, kidney, lipids, inflammation). We observed curvature; consequently, all models include age and age² to capture low-order nonlinearity with minimal complexity.

#### Cohort Characteristics, Sample Size, and Power

Each exposure-phenotype pair can be assessed in multiple surveys (1-10), and/or have multiple categorical levels (e.g., smoking is a categorical variable with 2 levels and a reference group).

We considered only phenotype-exposure pairs that had at least 500 participants across 2 or more surveys resulting in 278 unique phenotypes and 619 unique exposures with complete cases across all possible demographic covariates. The total number of associations possible to assess in 2, 3, 4, 5, 6, 7, 8, 9, and 10 independent surveys was 34,206, 21,451, 34,048, 9,011, 8,412, 1,702, 9,741, 999, and 1,455, respectively. To maximize power, for each association, we combined across 2 or more surveys, combining the sample weights as advised by the National Center for Health Statistics (see Methods).

The range of the sample sizes per association included 608 to 68,495 individuals. The median sample size was 7,464 and the interquartile range of 4,220 and 15,316. For a sample size of 608, the lowest R^2^ that can be identified was 0.05 with a power of 80% and a p-value threshold of 1×10^−6^. For the same power and p-value threshold, for sample size of 4,200, the lowest R^2^ that can be identified was 0.007; for a sample size of 7,464, the lowest R^2^ that can be identified was 0.004; for a sample size of 15,316, the lowest R^2^ that could be identified was 0.002; and for the maximum sample size (68,315), the lowest R^2^ that could be identified was 0.001.

### Statistical analysis

We cataloged a total of 322 real-valued phenotypes and 859 biomarkers or self-reported questionnaires responses that measure environmental, dietary, or behavioral exposures across all 10 surveys. Each exposure-phenotype pair can be assessed in multiple surveys (1-10), and/or, have multiple categorical levels (e.g., smoking is a categorical variable with 2 levels and a reference group). Across 322 phenotypes, 859 exposures, 10 surveys, and multiple variable levels, the total number of possible associations were 563,627. We give recommendations for ExWAS analytics in Table S9.

In our pipeline to maximize power and replication, we filtered for phenotype-exposures pairs that had at least 500-600 participants across at least 2 or more surveys (maximally a phenotype and exposure would appear in all 10 surveys), resulting in 305 unique phenotypes and 619 unique exposures. We calculated 119,643 associations between a phenotype (e.g., body mass index) and exposure (e.g., beta-carotene) pair. The total number of associations in 2, 3, 4, 5, 6, 7, 8, 9, 10 independent surveys was 34,206, 21,451, 34,048, 9,011, 8,412, 1,702, 9,741, 999, and 1,455. We used survey-weighted regression to associate phenotypes with exposures while adjusting for age, age^2^, sex, race/ethnicity, income, survey year, and education. We also performed associations with no adjustments (univariate) in each survey separately.

We associated *P* (total = 305) with every *E* (total = 651) in which overlapping samples were available across the surveys (total surveys = 10; 1999-2000, 2001-2002, 2003-2004, 2005-2006, 2007-2008, 2009-2010, 2011-2012, 2013-2014, 2015-2016, 2017-2018). An association could occur in 1, 2, 3, 4, 5, 6, 7, 8, 9, or 10 surveys. We used survey-weighted regression to accommodate complex survey design, including PSU, stratum, and subsample weights as input and implemented in ref^49^ (Extended Data Figure 1C-D). We considered two models to account for demographic-based confounding or stratification, a base model (as above) and a “demographic adjusted” model, which consists of covariates age, age^2^, race/ethnicity (Non-Hispanic White [reference], Non-Hispanic Black, Mexican American, Other Hispanic, and Other Ethnicity), household-to-income poverty ratio (ranging from 1-5), education (less than ninth grade, less than high school, high school graduate [reference], attended college, college graduate), and survey as a categorical variable. The summary statistics consists of the association sizes (beta coefficients), standard errors, p values, the sample size, surveys and R^2^ for the entire model.

For test statistic and p-value estimation, we report “robust” standard errors implemented in the survey package. We estimated the Bonferroni family-wise error rate and the Benjamini-Yekutieli false discovery rate (FDR)^50^ across all tests and primarily report findings that exceed either the family-wise error rate (Bonferroni). We also secondarily report the false discovery rate at thresholds less than 5%. We also estimate the “exposome inflation factor” ^51^.

We also estimated concordance of phenotype-exposure associations across independent survey samplings (Extended Data Figure 1DE). First, we calculated the variation of the phenotype-exposure associations across independent survey samples. For example, if a phenotype-exposure association was measured in 3 surveys, we estimated the association separately for each survey and calculated the variation of the phenotype-exposure correlation across the three surveys. We used a “unrestricted weighted least squares” meta-analytic approach ^52^ to estimate cross-survey standard error, and heterogeneity estimates (degree to which variation on association sizes can be explained across surveys, e.g. I^2^).

Separate and independent samplings of US NHANES by year (e.g., 1999-2000 or 2003-2004) provide an opportunity to estimate “replication” rates, or the times that a phenotype-exposure association has lower nominal significance level in greater than one survey sample. An association can occur in 2-10 surveys. For each scenario, we counted the number of times a phenotype-exposure association was achieved with p-value less than 0.05; for example, for a phenotype-exposure association that can be assessed in 5 surveys, that phenotype-exposure association can be significant up to 5 times.

#### Exposure-Exposure Correlation Globes

We measured the partial Pearson correlation between 619 exposures with pair-wise complete data, a total of 200k correlations. The partial correlation is the correlation between exposures, adjusted by age, age^2^, income to poverty threshold, education, and ethnicity. To visualize an “exposome globe” as an example, we queried the correlation database for correlations among randomly selected exposures and exposures associated with Body Mass Index or Glycated Hemoglobin. We filtered to only visualize correlations that were greater than 0.25. We visualized the globe using the *igraph* R package ^53^.

#### Shared associations between exposures and phenotypes

We defined the “shared” associational architecture between pairs of exposures (e.g., phthalate and heavy metal) or phenotypes (e.g., body mass index and glucose) by the similarity of their phenotype-exposure associations. Specifically, for a given exposure, there is an array of associations estimated for *X* number of phenotypes (e.g., a row in the Atlas, Figure 3). We estimate the correlation distance between a pair of exposures. For example, if a pair of exposures (e.g., E1 and E2), have the same association for each n phenotypes (e.g., P1… Pn), their association architecture is equal and the correlation will be 1.

#### Estimating the Aggregate Contributions of Genetic and Exposomic Factors to Phenotypes

To compare the variance explained by exposomic and genetic factors across 29 phenotypes, we constructed and evaluated multivariate models using a three-step workflow. First, for exposome-based modeling, we identified exposures significantly associated (FDR-adjusted p-value < 0.05) with each phenotype using univariate regressions. Among these, we selected up to the top 20 exposures based on their univariate R² contributions. We multiple imputed missing data using Multiple Imputation with Chained Equations (MICE) ^54^, generating 10 imputed datasets via predictive mean matching. For each phenotype, we fit two linear regression models: a baseline model incorporating demographic covariates (age, sex, ethnicity, education), and an expanded model that additionally included the selected exposures. The incremental R² attributable to exposures was calculated as the difference in variance explained between these two models.

Second, for genetic modeling, we leveraged previously published genome-wide association study (GWAS) summary statistics derived from approximately one million genetic variants (UK Biobank) across 125 quantitative phenotypes of anthropometry and biomarkers. 29 of these 125 phenotypes (23%) overlapped with those queried in this investigation. Genetic contributions (R²) to each phenotype were obtained from quantitative trait GWAS models that included age, sex, genotyping array, and principal components as covariates. The genetic R² was defined as the incremental variance explained by the genetic model beyond these baseline covariates. The median genetic R^2^ across all 125 phenotypes queried as reported in ref was 7% (interquartile range of .4% and 9%; max R^2^ of 36%).

We directly compared exposome-based and genetic-based R² values across the 29 overlapping phenotypes (Table S4). We selected phenotypes based on the availability of matched phenotypes data from NHANES and the UK Biobank. This allowed assessment of the relative explanatory power of exposomic versus genetic factors for each phenotype, critically informing the conclusions about the comparative utility of exposomic data relative to genetic predictors.

#### Concordance of associations with different types of measurements of the same exposure

Some variables are assessed at different times (part of a 2-day interview) or sample matrices (e.g., in both urine and blood). We examined the concordance of phenotype-exposure associations across these samplings (e.g., day 1 exposure vs. day 2, or blood vs. urine measure). For self-reported nutrients, we compared the phenotype-exposure associations for day 1 vs. day 2 Number of dietary supplements reported, Calcium, Carbohydrate, Copper, Folic acid, Folate, DFE, Iron, Energy, Lycopene, Lutein + zeaxanthin, Magnesium, Niacin, Phosphorus, Potassium, Selenium, Thiamin (Vitamin B1), Vitamin B12, Riboflavin (Vitamin B2), Vitamin B6, Vitamin C,Vitamin D (D2 + D3), Vitamin K, Zinc, Total fat, Iodine, and Total sugars. We also compared the phenotype-exposure associations between self-reported measures and their biomarker counterparts, including alpha carotene, vitamin b12, cis beta-carotene, cryptoxanthin, retinol, trans-beta-carotene, and vitamin D. Last, we compared phenotype-exposure associations between blood-based and urine-based indicators, including mercury, cadmium, and cotinine.

## Data Availability

The full cohort database can be found here at doi: 10.6084/m9.figshare.29182196. The full summary statistics can be found at doi: 10.6084/m9.figshare.29186171. NHANES data are publicly available: https://wwwn.cdc.gov/nchs/nhanes/default.aspx. All results from the PE-WAS can be visualized, downloaded, and browsed online: https://pe.exposomeatlas.com Specifically, users may query an *ExWAS* for a specific phenotype and examine phenotypic specific associations, R^2^, and exposome correlation globes.

## Code Availability

All code is available as a R analytics package developed under an MIT license here: https://github.com/chiragjp/nhanespewas. Analytic code was written under R version 4.4.2 (R versions supported include >= 4.0. The libraries required for the package include DBI,RSQLite, dplyr, tidyr, survey, stats, tibble, broom, rlang, tidyselect, logger, purrr, stringr, ggplot.

## Declaration on Ethics and Consent

This study was deemed “not human subjects” research by Harvard Institutional Review Board (IRB): IRB24-1004. CDC NHANES participants have previously consented their data for use in research, and the protocol has been approved by the National Centers for Health Statistics (NCHS) ethics board: https://www.cdc.gov/nchs/nhanes/about/erb.html

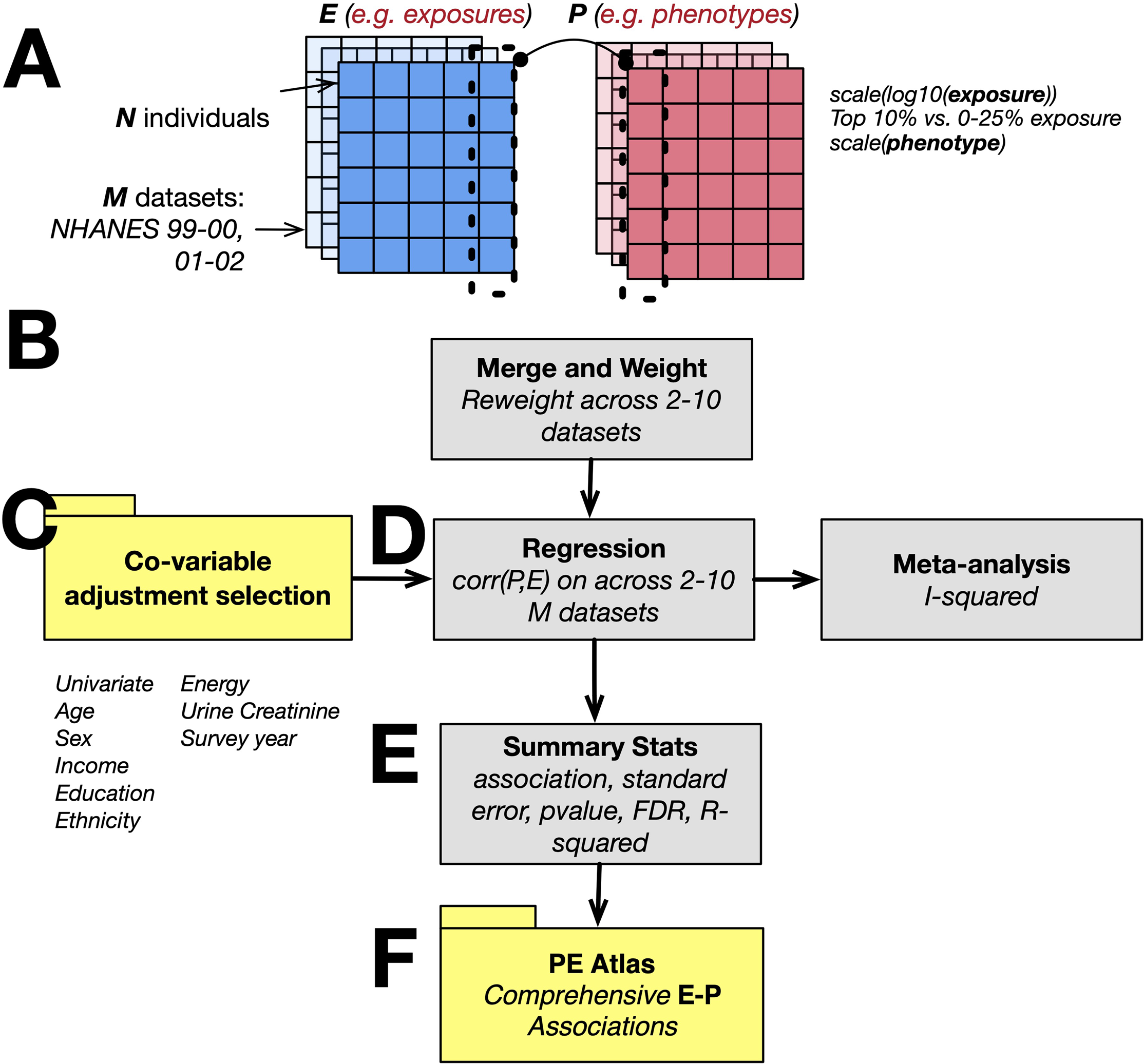

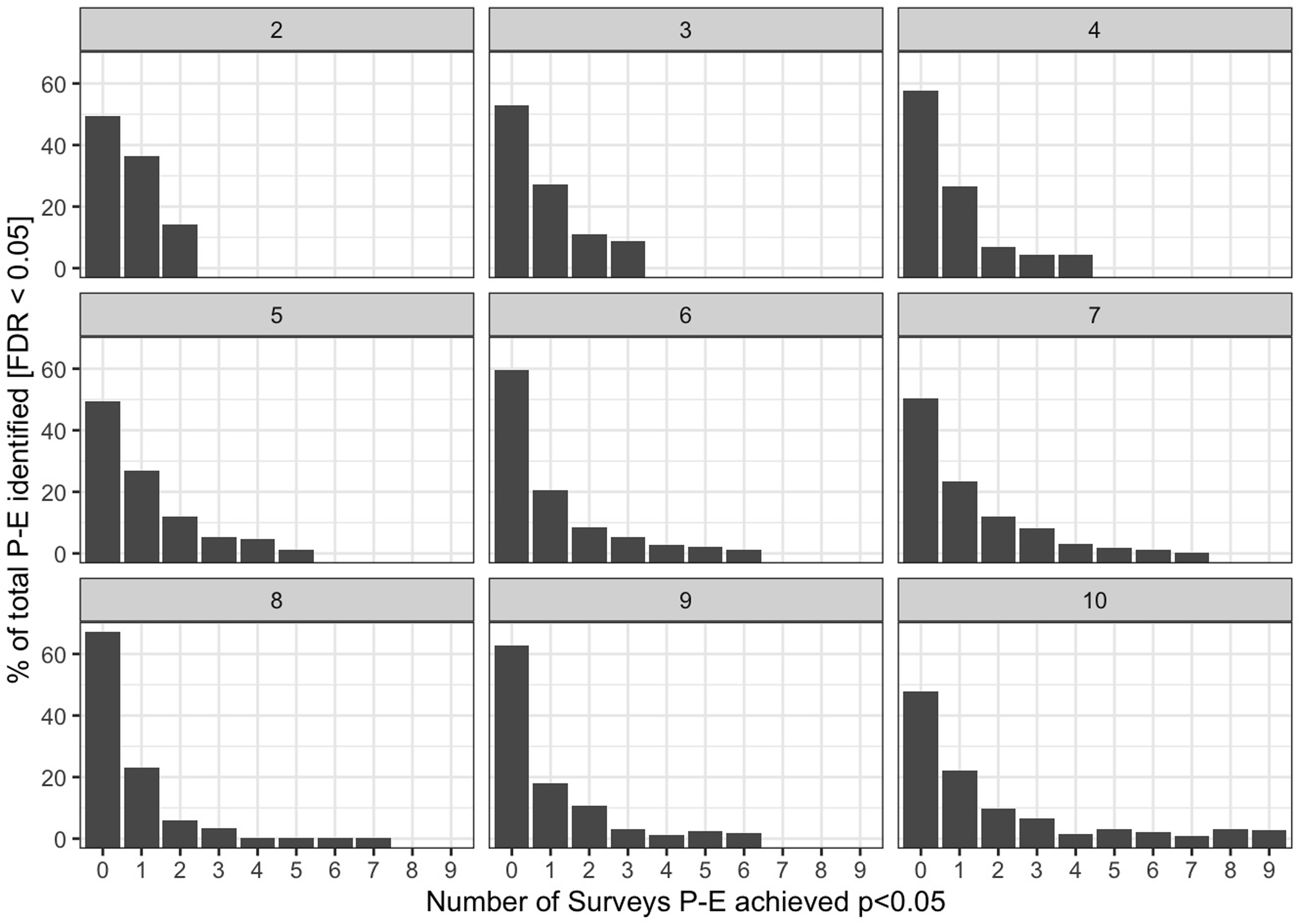

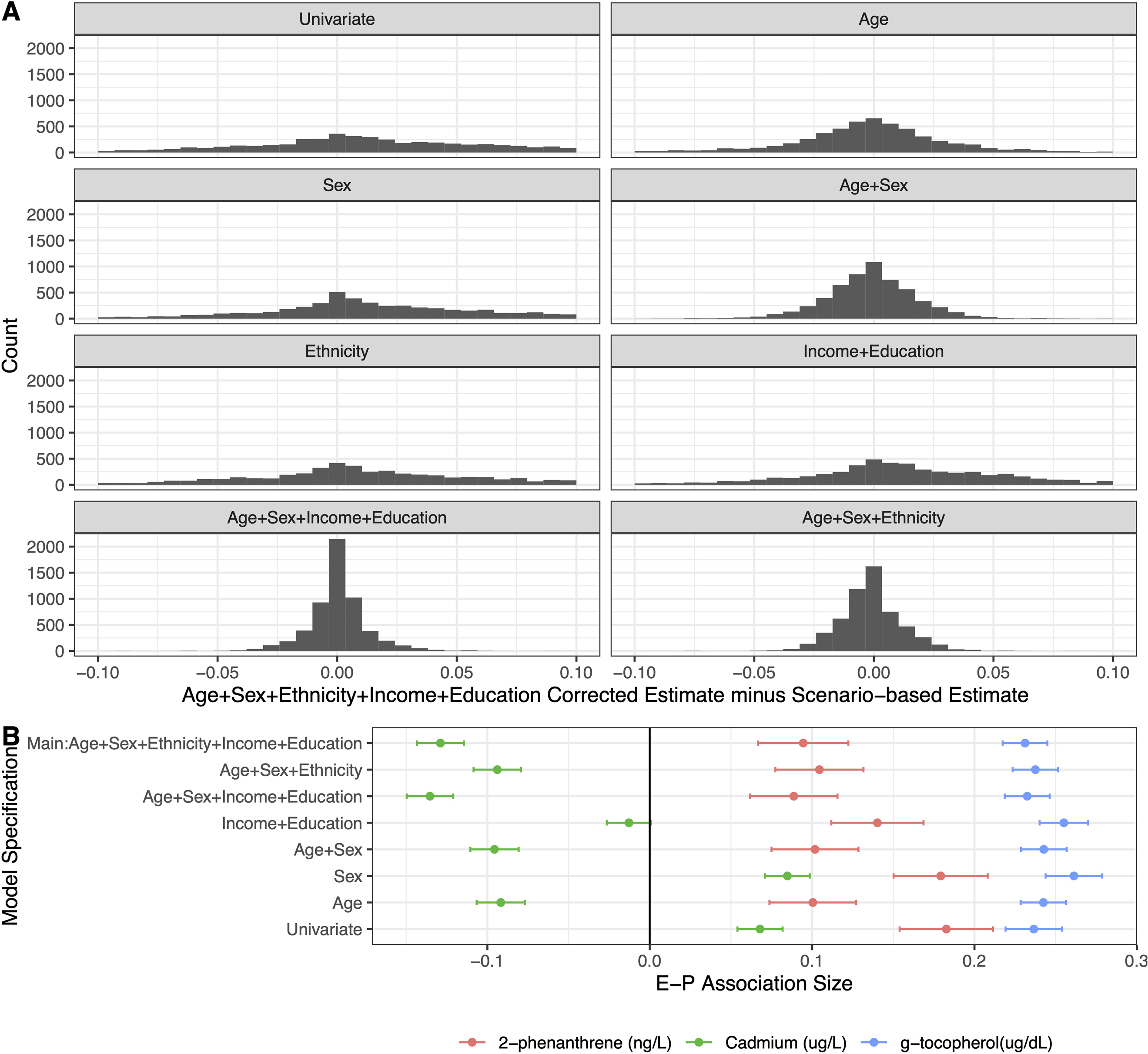

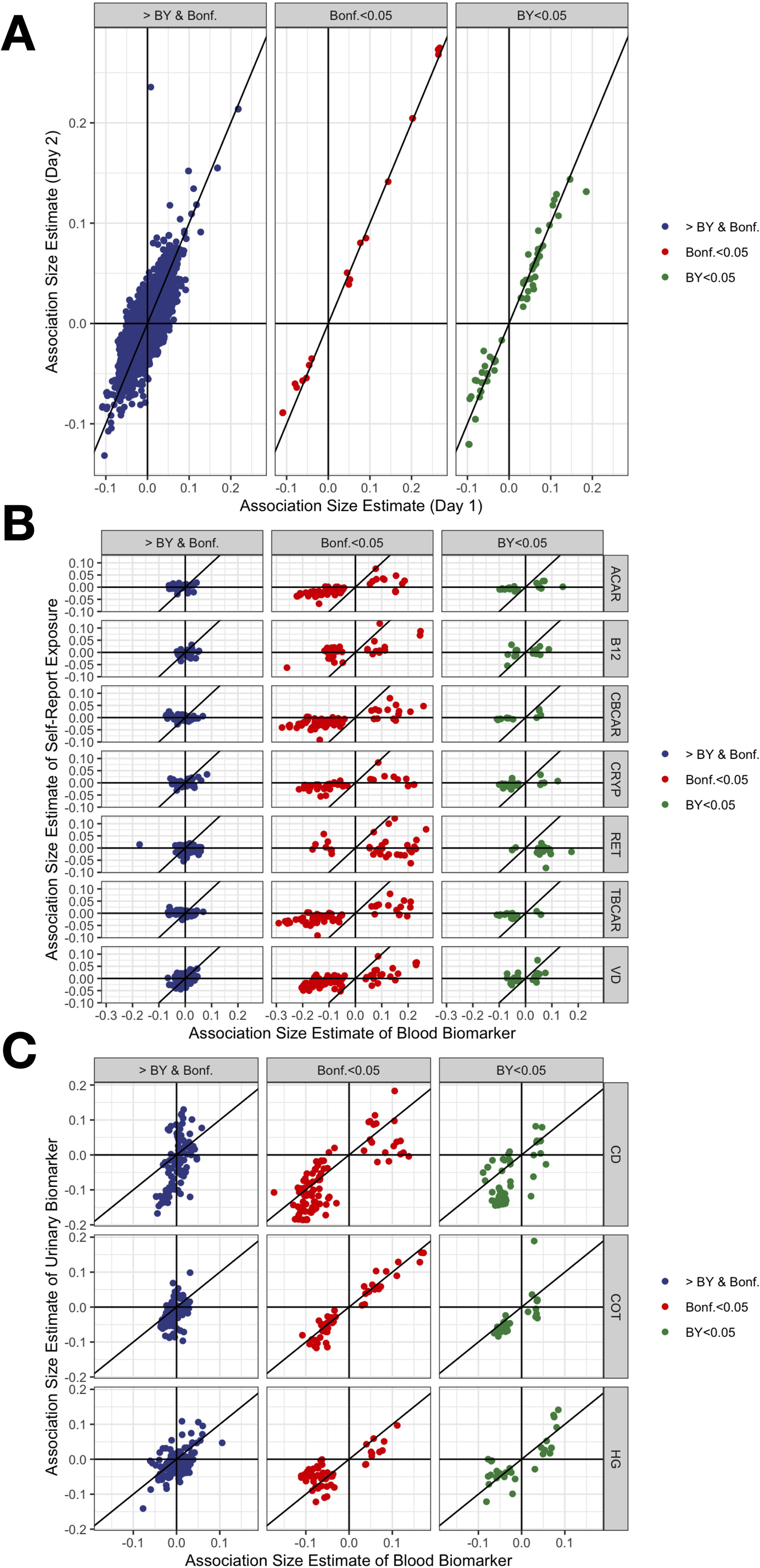

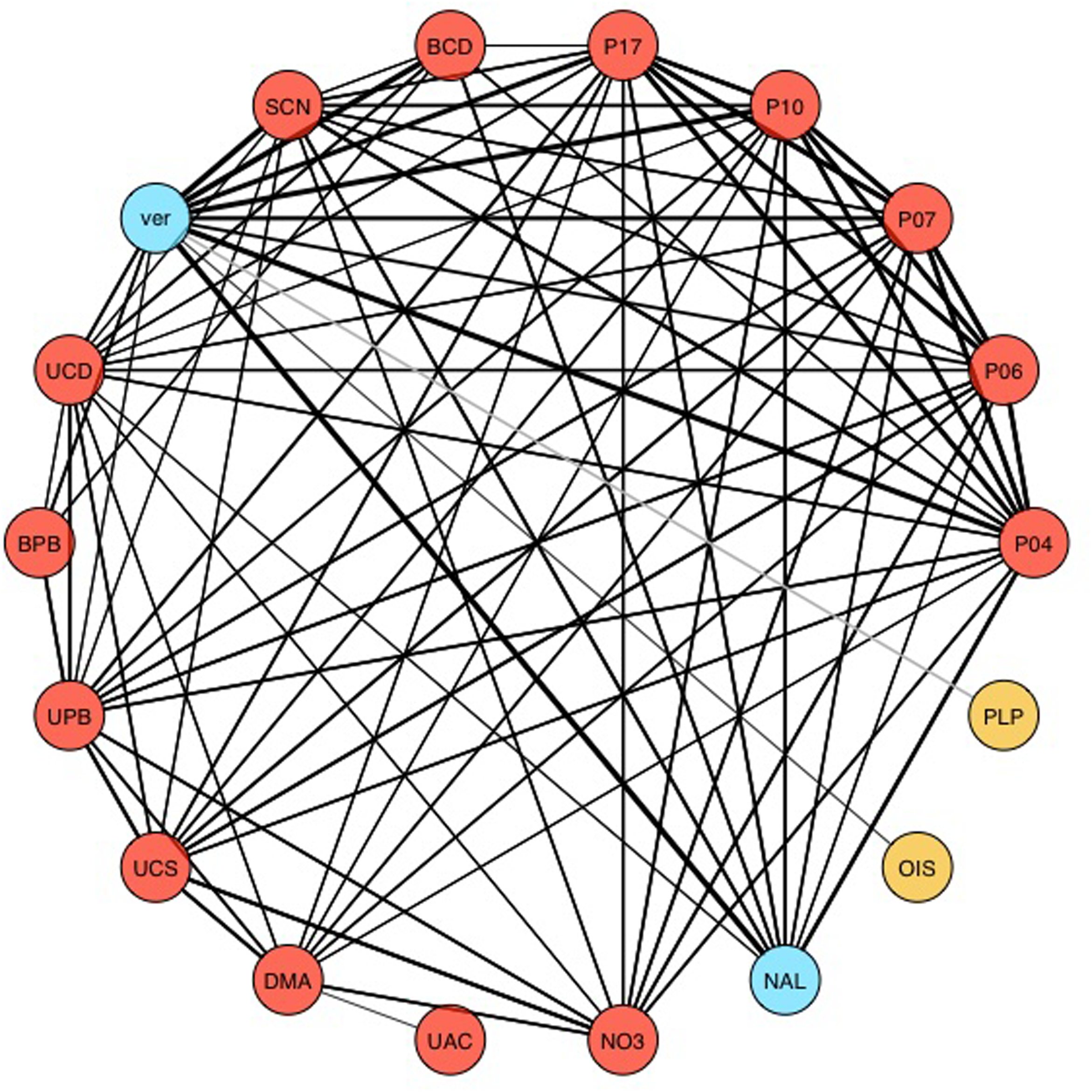

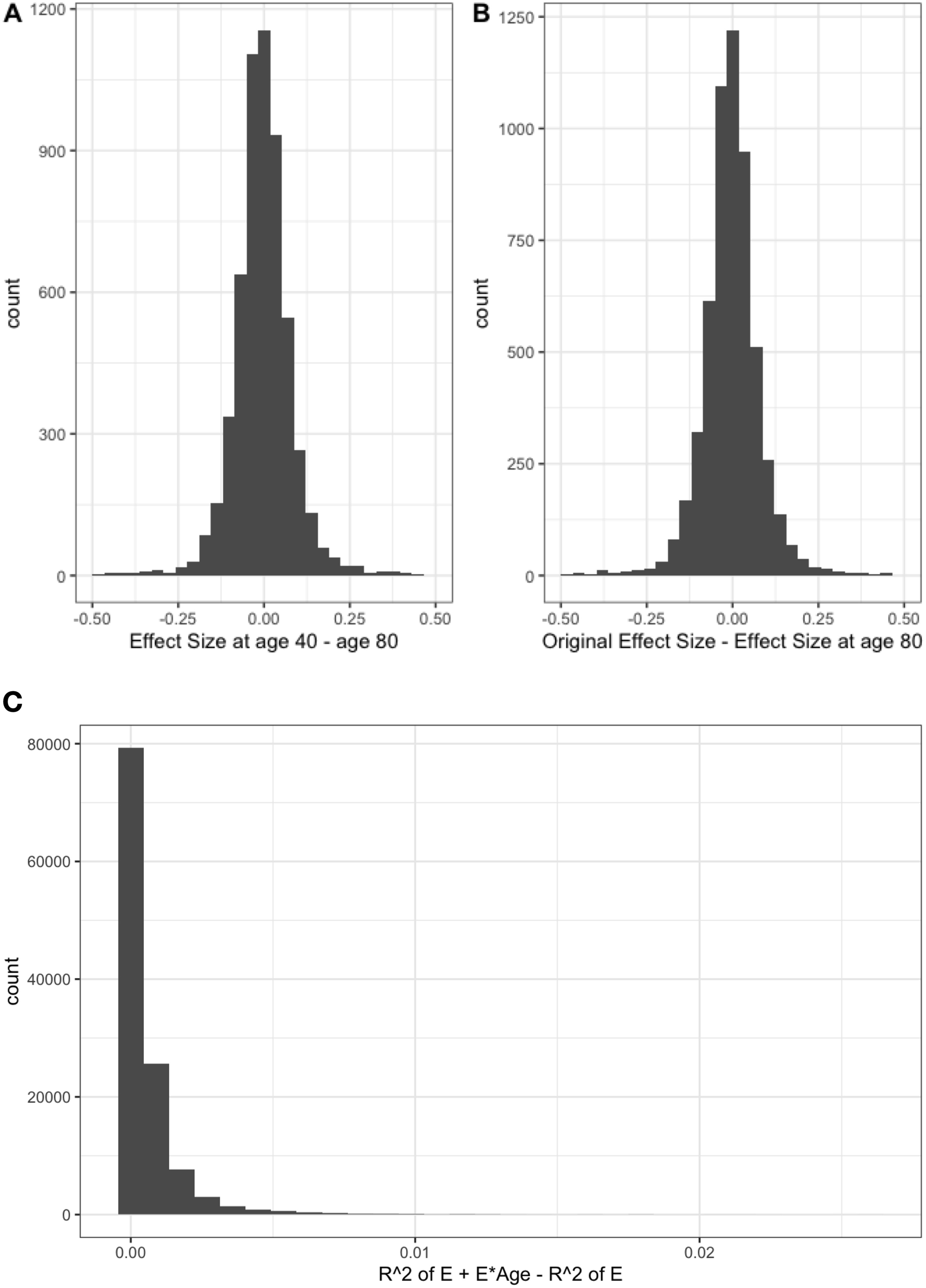

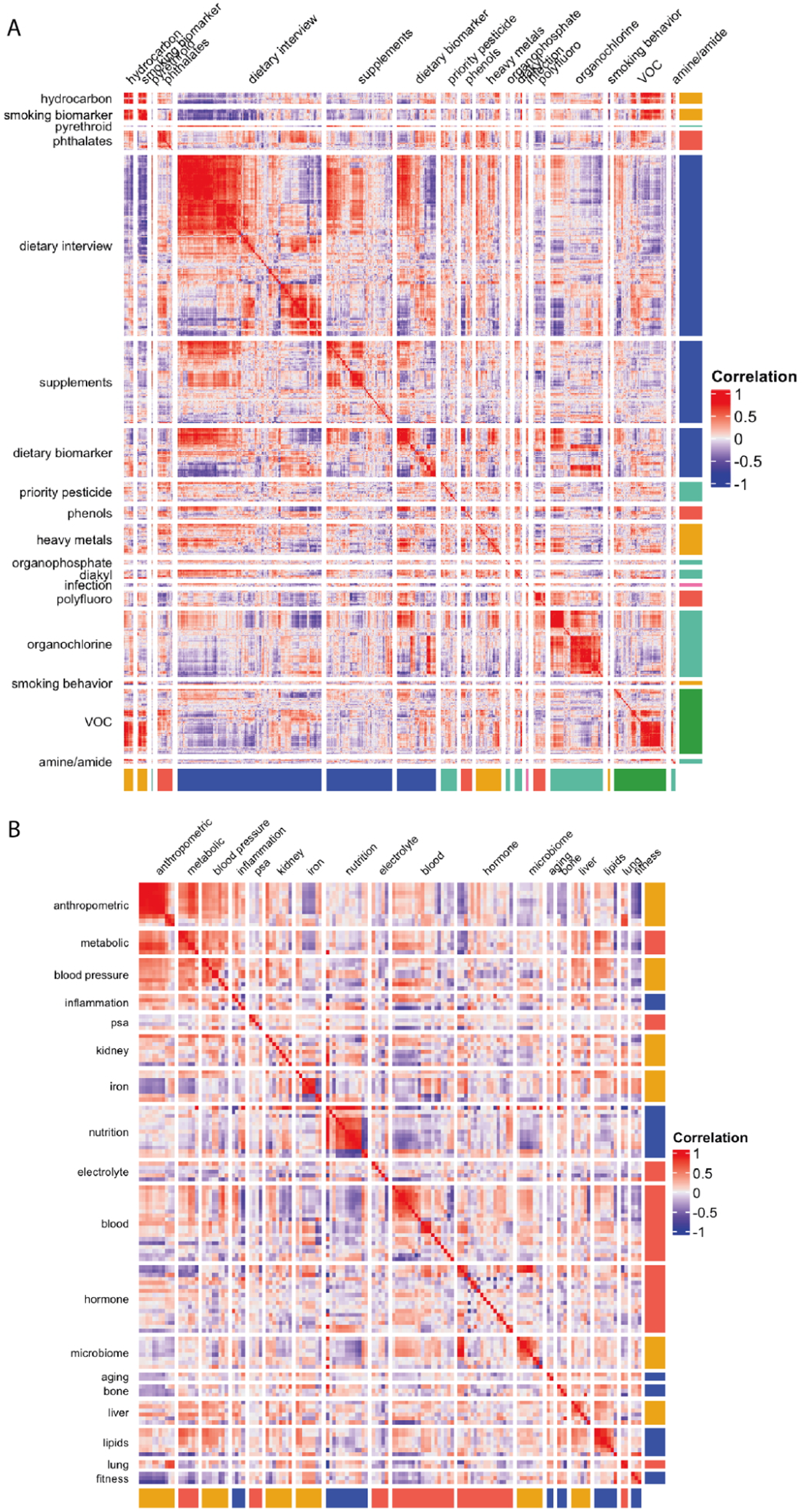

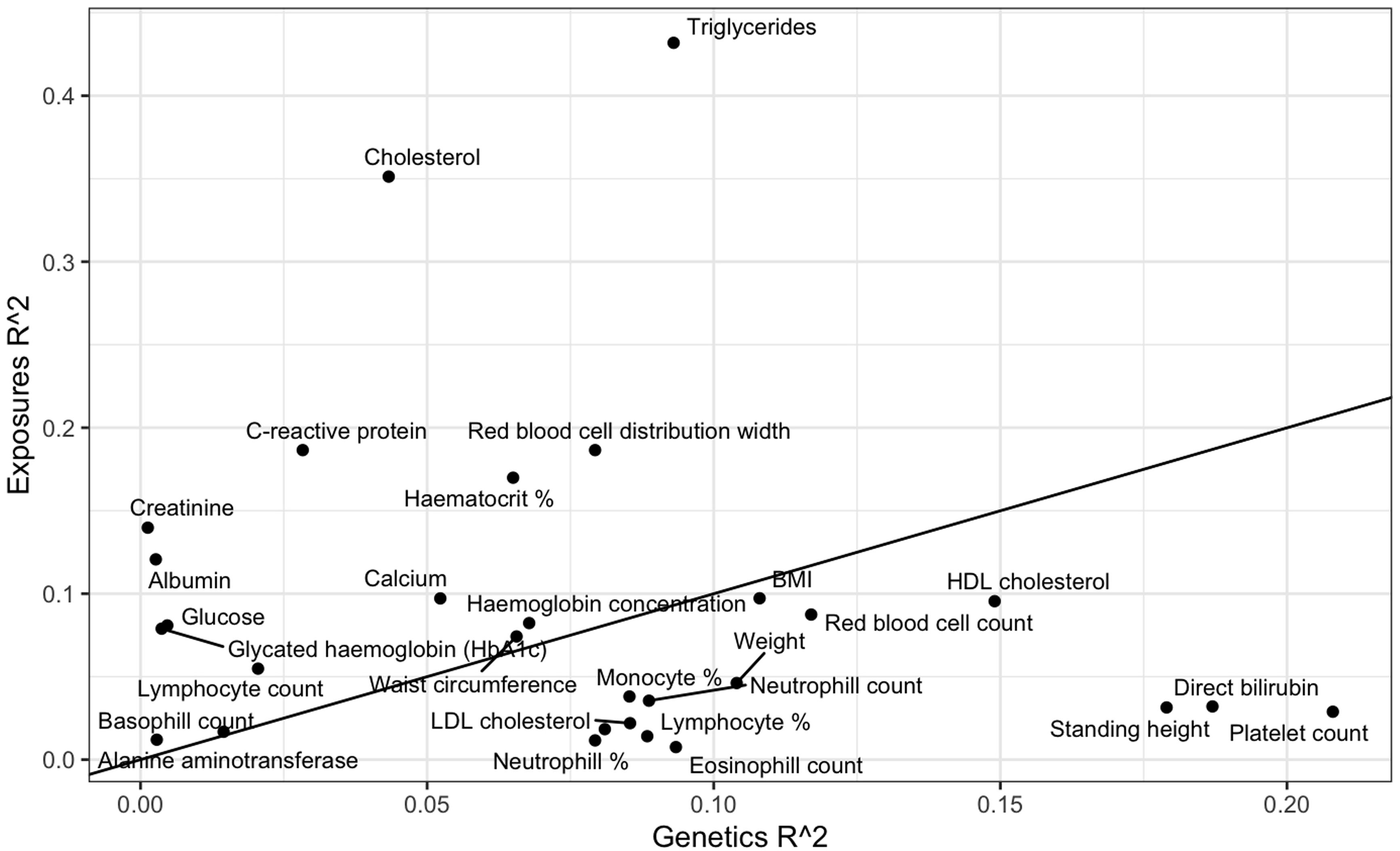

